# Barriers to contraceptive use in humanitarian settings: Experiences of South Sudan refugee women living in Adjumani district, Uganda; An exploratory design using qualitative method

**DOI:** 10.1101/2022.11.23.22282662

**Authors:** Roselline Achola, Lynn Atuyambe, Elizabeth Nabiwemba, Mathew Nyashanu, Christopher Garimoi Orach

## Abstract

**Introduction:** Family Planning (FP) is a life-saving, empowering and cost-effective interventions for women and girls. Access to FP is still challenging to female refugees due to several barriers including language, low educational level, lack of information, influence by significant others, limited income, desire to replace lost family members, moral values, cultural and religious norms as well as personal experience with contraceptives side effects. This study explored barriers to contraceptive use among South Sudan refugee women living in Adjumani district, Uganda.

**Methods:** An exploratory design using qualitative method was employed involving women between the ages of 15-49 years. Purposive sampling was employed to select participants for Focus Group Discussions (FDGs) and In-depth Interviews (IDIs). Ten FDGs were conducted, each consisted of 6 to 8 participants. Two groups of females (15-19 and 20+ years) were reached. Twenty-seven IDIs were conducted with two similar categories as above. The In-depth interview and focus group discussion guides were used to collect data. The interviews were recorded verbatim and transcribed before being translated back to English from the local language. Audio recordings from the FGDs and IDIs were labelled, transcribed verbatim and translated into English. Deductive, team-based coding was implemented for codebook development, Transcripts were entered, and data coded using Atlas ti version 14. Data was analyzed using content analysis to produce the final outputs for the study.

**Results:** The study found that several challenges to contraception use included gender dynamics, socially constructed myths on contraceptive use, cultural norms related to contraceptive use, limited knowledge about contraceptive use, men’s negative attitudes, antagonism of contraceptive use by leaders and reprisal of women who use contraception.

**Conclusion:** The study concluded that there is need for community strategies to break down the barriers to contraception utilization among refugee women. Such strategies should include men and women alongside gatekeepers to enhance sustainability.

## Introduction

The last twenty-five years have witnessed massive displacement of people due to political upheavals and natural disasters [1]. This has precipitated the establishment of refugee settlements in many countries.

More so, the displacement and subsequent establishment of refugee settlements is common in low and middle income countries (LMICs) whose infrastructure and capacity for support is poor. The refugee population are affected by an array of problems ranging from economic, socio and cultural due to living in new environments with poor supporting systems [2]. In addition, family planning is also an urgent need currently affecting refugee communities considering that family size has a direct impact on family well-being and support.

Family Planning is a life-saving, empowering and cost-effective interventions for women and girls. However, it remains underfunded with limited prioritization in humanitarian responses with some restrictions by host government on access to services [3]. When immigrants take refuge in other countries, demand for education, health services, infrastructure, natural resources, food, land and security concerns are considered key [4] with lesser prioritization of family planning as a key pilar to improved quality of life for refugee women of reproductive age.

Global standard for Sexual Reproductive Health response in acute emergencies identified prevention of unintended pregnancies as one of the six objectives under Minimum Initial Services Package for Sexual Reproductive Health programming (World Health Organization, 2020). But many women and girls who would like to access Family planning services have challenges with family decision to allow them to utilize the services. Family planning, according to World Health Organisation report, 2020 was defined as a voluntary and informed decision by an individual or couple on the number of children to have and when to have them. It is characterized by the use of contraceptive methods that include: female and male sterilization, condoms, injectable, oral pills, implants, Intra-Uterine Devices (IUD) of both hormonal and Copper T, Lactation Amenorrhea Method (LAM) among others [5]. Additionally, Family planning also includes traditional methods comprising of withdrawal, periodic abstinence, basal body temperature among others [6].

The level of accessibility of FP services by refugees in the host country is often affected by several barriers including language, low educational level, lack of information, influence by significant others, limited income, desire to replace lost family members, moral values, certain taboos, cultural norms, religious impediments and personal experience with contraceptives side effects [7];[8]. In addition, socio-cultural preference and unacceptability of contraception also pose significant barriers to making decision to use FP [9]. This is more prevalent in many patriarchal societies where men are viewed as the custodians of the community [10].

With global displacement of over 89 million people by war, violence and other forms of human rights abuses [11], refugee populations have continued to face vulnerabilities characterized by living in volatile environment and among the people of different cultural norms that they are not accustomed to. Uganda is the 3^rd^ largest refugee-hosting country in the world and currently host over 1.5 million refugees from neighbouring countries and beyond [11].

These refugees are spread all over the country with a high concentration in the west Nile and western regions. Over ninety-four percent are living in settlements established in 12 of Uganda’s 135 districts and 6% in Kampala. Of these, (60.5%, n = 964,960) come from South Sudan and 29.3%, n = 467,004) come from the Democratic Republic of Congo and others 10.2%, n = 163,441) from Somalia, Burundi, Rwanda, Eritrea, Ethiopia and Sudan [11]. Of the total number of refugees who come from South sudan, about 214,453 live in settlements in Adjumani district (OPM, 2020). Over 80% of the refugees are women and children. These vulnerable groups need cost effective interventions for improved Sexual Reproductive Health and Right (SRHR). Although it is known that a woman’s ability to space and limit her pregnancies has a direct impact on her health and well-being, contraceptive use is still low at 34% in the least developed countries.

South Sudan is one of the countries whose modern contraceptive prevalence rate is very low at 2.7% with unmet need for FP at 30.8% with maternal mortality ratio of 789/100,000 live births [12]. With the health challenges back home in South Sudan where infrastructure was destroyed during the war with limited economic opportunities, women still face challenges of limited access and male dominance over their Sexual Reproductive Health Rights. The women’s position is a subordinate one with husbands being sole decision makers including determining the number of children to be born [13]. Thus, women are expected to produce as many children as possible in return for the cows paid as dowry to their parents [14]. The consequences women face when found using modern FP methods include beating and even separation or divorce. This exposes women to the barriers to access to FP information and services leading to unintended pregnancies. Considering all the above assertions this study aimed at exploring the barriers to contraceptive use in humanitarian settings: challenges and opportunities for South Sudan refugee women living in Adjumani district, Uganda.

## Methodology

### 2.0 Methods

#### Study design

An exploratory design using qualitative method was employed to explore the barriers to contraceptive use in humanitarian settings for South Sudan refugee women living in the three settlements in Adjumani district, Uganda. This method is important in understanding the problem deeper as opposed to finding a final solution to the problem [15]. Furthermore, the study is guided by the ecological perspective theory which describes how different levels interact differently to cause an outcome which in this case is the decision making for FP use among refugees. [16]. Mcleroy added; I quote, ***“****An ecological perspective on health, emphasizes both individual and contextual systems and the interdependent relations between the two”* [16]. The researchers adopted this theory because decision making on health behavior is central to this study while behaviour is affected by several levels of influence indicated in the ecological theory (See fig 1 below). Ecological perspective theory describes how behavior shapes and is also shaped by the environment in which individuals live [16]. It describes eight theories and models that explain individual, interpersonal, community behaviour and offers approaches to solving problems [17] [18]; [19]. The levels of influence include: (1) intrapersonal/individual factors (2) interpersonal factors (3) community factors (4) institutional/organizational factors (5) public policy factors. This explains how the different levels of influence interact towards population health, which in this case is the use of Family Planning for improved health. **(Fig 1)**

**Figure 1:**
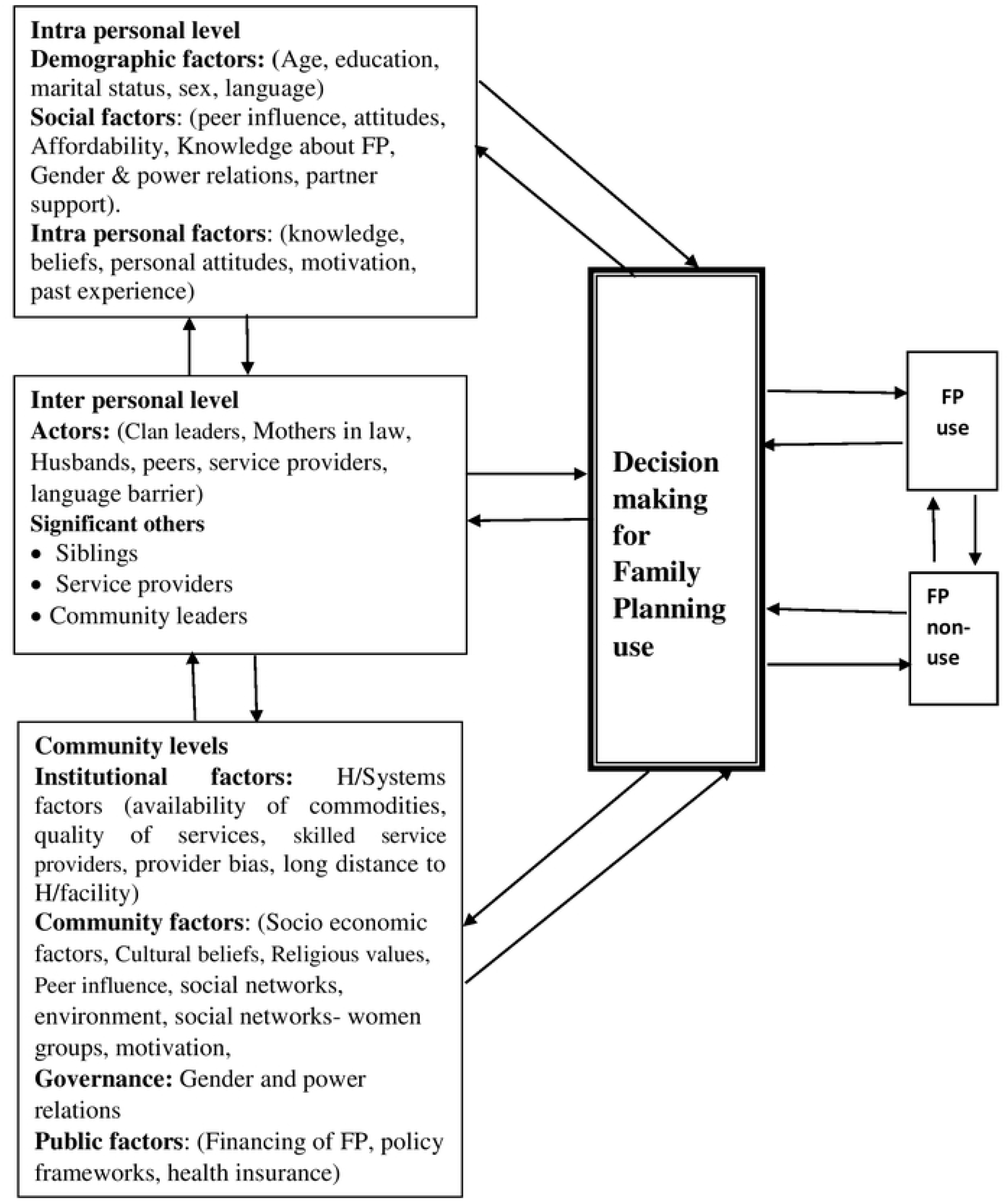
Different interactions between different levels to cause an outcome of decision making for Family Planning use. Adopted from Ecological perspective.

#### Study population and recruitment

This study was conducted in Adjumani district in the three settlements of Pagirinya, Nyumanzi and Mirieyi. The study participants were women of reproductive age (15-49 years), both users and non-users of contraceptives. The study participants mainly spoke Madi and Arabic. In this study, purposeful sampling approach [20] was used to select participants for FGDs and IDIs with support of community leaders and service providers from the health facilities. Ten FDGs were conducted overall. Each focus group consisted of 6 to 8 participants. The groups of females reached were of ages 15-19 and above 20 years. Twenty-seven IDIs were conducted with similar categories of participants.

#### Piloting the interview schedules

Whereas the research assistants were knowledgeable and familiar with study area and the local languages, they were trained on the basics of qualitative research and taken through the tools. Having identified the 3 settlements with their host communities, the pre – testing of the tools was done in Agojo settlement and its host community in the same district. Purposive sampling was used to recruit participants who took part int the FGDs and IDIs. The research participants were asked about the clarity of the questions which they confirmed to be clear, thus the researchers adopted the interview guides without much alteration.

### Data collection

The interview schedule was translated into the local languages to facilitate understanding of the questions by the research participants. Research assistance were utilized to collect data from the research participants. The Research assistants who were recruited to help with data collection were able to speak the local languages which included Dinka, Madi and Arabic. They were trained on qualitative research methods and the use of tools by taking them for field pre-testing in similar settings. Data was collected using two interview guides. The In-depth interview and focus group discussion guides. The interviews were recorded verbatim and transcribed before being translated back to English from the local language.

### Data Analysis

Audio recordings from the FGDs and IDIs were labelled, transcribed verbatim, translated into English by experienced research assistants who were fluent in Madi and Arabic as the main languages and the recordings stored. Deductive, team-based coding was implemented for codebook development, Transcripts were entered and data coded using Atlas version 14. Data was analyzed using content analysis. To ensure reliability, coding was done by three research experts led by the Principle Investigator. They read the transcripts, discussed emerging issues and agreed on common themes. The team selected two transcripts, each one read through, assigned different meaning to each response and developed a codebook following Braun and Clarke procedure [21]. On completion, they met on zoom and discussed the descriptive codes to come up with a unified code book. While coding, there were interactive discussions arising from the discrepancies and disagreements that were resolved by having same meaning of the codes before moving to next step.

#### Ethics statement

This study was approved by Higher Ethical Degree Review Committee of Makerere University School of Public Health (25^th^ March 2021) and the Uganda National Council of Science and Technology. Written permission was also obtained from Office of the Prime Minister (19^th^ April, 2021), Kampala and in Adjumani Sub Office. All the research participants read the information sheet and signed a consent form, which granted them the right to withdraw from the study at any time without giving reasons.

## Results

Following data analysis, the study found that the challenges to contraception use included gender dynamics, socially constructed misconceptions on contraceptive use, cultural norms related to contraceptive use, limited knowledge about contraceptive use, men’s negative attitudes, the antagonism of contraceptive use by leaders and The reprisal following contraceptive use.

### 1. Gender dynamics

#### 1.1 Joint decision by couple against contraceptive use

The research participants in the refugee settings viewed contraceptive use as a barrier to getting the many cows in form of bride price that comes with a girl child. Families that produce many girls are assured of big gains in form of dowry. Therefore, participants reaffirmed that couples do agree jointly not to use contraception and have as many girls as possible to reap the benefits of dowry.

> “…*the problem is that every man tells the wife about the need for many cows and the easiest way is give birth to more girls which your contraceptives will deter them from achieving…”* (IDI_Female_20+_Adjumani_Nyumanzi_ settlement).

> *“…Back home in South Sudan, we even don’t use contraceptives because what we want is to give birth to many children. This is the reason why we don’t want to know more about your family planning. In our tribes, we face poverty due to lack of dowry, so we need more girls to get out of poverty…”* (FGD_female 20+ _Adjumani_Mirieyi_settlement - Dinka only).

#### 1.2 Limited women empowerment

The research participants acknowledged that women who are less empowered are unable to convince their husbands about the intention to use contraceptives. They reported that personal decision is only possible for women who are economically empowered and are able to sustain themselves and provide basic needs should the husband discover that they are using any contraceptives.

> *“…you see these men of ours they don’t want this family planning, they say it will damage people and its dangerous for women to use even though you share with them the benefits of Family Planning, for them they will disagree and even become furious…”* (IDI _Female 20+_ Adjumani_Pagirinya Settlement)
>
> “…*if you are very weak the husband will chase you away from that home…”* (IDI_ female_ 20+_Pagirinya_Adjumani_settlement).

#### 1.3 Patriarchal dominance of men

The research participants reported influence of men in making decision on contraceptive use. They further reported that women find it hard to make decision to start on a contraceptive without husband involvement apart from doing it stealthily which also has negative consequences. They acknowledged that men were the custodians of decision making in their communities and key actors in determining family size and family planning use.

> “…*Because my husband was aware that my mother made decision for me to start on family planning methods without his consent, he said that if I fall sick in his house, I should go to my mother who should take care of me moving forward*.…” IDI _Female 20+_ Adjumani_Pagirinya_Settlement
>
> *“…I witnessed a sad moment when my neighbor made her decision to go to hospital and was given this method of inserting on the arm, how do you call it? Implant yeah. But when this woman got sick, while the husband tried to support her, he felt the implant under her arm. He immediately rushed to hospital with her and commanded that the implant be removed off the wife’s arm as he was looking*…” IDI
>
> _Female_20+_Adjumani_Pagirinya_Settlement.

#### 1.4 Mistaking contraceptive use for promiscuity

The research participants reported that many husbands and parents of young women believed that women who use contraception are promiscuous. Furthermore, they alleged that the community views contraceptive use as a cover up for promiscuity, thus strongly oppose its use by both women and girls.

> *“…yes, it is now more of a culture in the Dinka tribe, that women who are using contraception when the community gets to know, they will be labeled women who don’t want to produce because they are prostitutes. This has discouraged more women from using contraceptives even when they have genuine reasons…”* **(FGD-Female 20_Adjumani_ Pagirinya _settlement)**
>
> *Most parents don’t want their young girls to use contraceptives, saying the girls want to become prostitutes by covering up with prevention of pregnancy*. **IDI-Female 20+_Adjumani_ Pagirinya _settlement**

### 2. Socially constructed myths and misconceptions about contraceptive use

Research participants reported that in their communities, people have negative perceptions about contraceptive use. They reported that contraceptives cause cancer, infertility, fibroids, burn the ovary, damage the female eggs and even lead to giving birth to deformed child. Furthermore, they claimed that children born with deformities are as a result of using a contraception. They also reported that perceived beliefs that are related to myths and misconceptions demoralize most women who might be considering using contraceptives.

> “… *I think if women are not determined due to the myths that go around about various contraceptive methods, no one would probably use given the negative forces that come your way. People say that Depo causes cancer, fibroids among others*…*”* (IDI_Female 20+_Mirieyi_settlement)
>
> Research participants added that some parents claim that Family Planning use among young people is very bad. They reported that when girls who are not yet married use contraceptives, they may not have children for the rest of their lives in addition to deformed babies.
>
> “…*They also say that they may have babies with missing body parts. So, any abnormality is associated with use of modern Family Planning. Even those who don’t produce after marriage are labelled as having used Family Planning at early age*. (IDI-Female 20+_Adjumani_Pagirinya_refugees)

#### Cultural norms related to contraceptive use

Most of the research participants reported that women are married to have as many children as they can until when they naturally reach menopause. Thus, failure to have the desired number of children can tantamount to disciplinary action against the woman because it contravenes the cultural beliefs and norms on big family size.

> *it’s there, cultural belief that husbands would tell their wives that I brought you to my home here to produce children and you cannot stop before your age*. IDI_female_20+_Adjumani_Pagirinya_settlement.
>
> *“…even now you just see people they are fearing family planning and if you go in communities, the fellow women just laugh at you saying this one doesn’t want to produce. what’s wrong with this woman*… *laughing even if you talk to them they continue looking at you as one who doesn’t want to produce…”* IDI_ Female-_20+_Adjumani_Nyumanzi_Settlement.

### 3. Limited knowledge about contraceptive methods

The participants reported that contraceptive information is not widely known by the refugee communities. Additionally, they acknowledged that there was lack of health promotion messages that promoted family planning back home in South Sudan.

> *“…I don’t use family planning because I don’t know more about it. No one comes to talk to us in the settlements here about the use of family planning…”* (FGD_Women_20+_Adjumani_Mirieyi_Settlement_DINKA)
>
> *“…things would be different because here in Adjumani, one can mention anything about contraceptive which is not the same case back home in South Sudan…”* (Female FGD_20+_Adjumani_Pagirinya_ settlement).

### 4. Men’s negative attitudes about contraceptives

The research participants reported that their main barrier to use contraception is due to negative attitudes of men about family planning. They added that men think that their women will stop producing.

> *“…you see these men of ours don’t want this family planning, they say it will damage people and its dangerous for women to use even though you share with them the benefits of Family Planning, for them they will disagree and disagree and start quarrelling*…” **IDI _Female20+_Adumani_Pagirinya_Settlement**.
>
> Surprisingly, research participants added that this negative attitude by men have also influenced the way women now perceive family planning. They reported that women also speak negatively about use of contraception.
>
> *“…But for women who listen to their husbands have also developed negative attitudes about family planning. They discourage other women, saying that family planning method is not good because women may give birth to a child who is dormant (with mental retardation), lame (with deformity) …”* IDI _Female_20+_Adjumani_Pagirinya_Settlement.

### 5. Antagonism against contraceptive use by leaders and men

The research participants reported that, there was a deliberate effort by the community leaders to antagonize family planning programs within the settlements. They continuously advocate for many children to build a stronger and bigger society, claiming that conflict back home disentangled their families that must be rebuilt with new generation. The participants added that their leaders have a fear of losing the entire generation and is the reason why they antagonize family planning programs for refugees. The research participants also reported that children are viewed as source of prestige and security. Therefore, more children are needed.

> *“…it’s common in Madi or any culture among us refugees that we are interested in replacing our lost people. Therefore, we need to give birth to many children who will be going to occupy our land back in South Sudan. Even if you go back there, that land is now open no people. Thy are just advising women to produce as many children as they can…”* (IDI female_ 20+_ Adjumani_Pagirinya_Settlement).
>
> *“…South Sudane man with a woman who has given birth to less than five children and was not conceiving again, the clan leaders force the man to marry another wife because more children are needed*…*”* **IDI female 20 and above Pagirinya settlement**

### 6. The reprisal following contraceptive use

The research participants reported that women who went ahead to use contraceptives when their husbands and other family members had rejected the decision, received some reprisals. They asserted that it was safer when the husbands and family members remained ignorant about their contraceptive decision.

> *“… Some women are neglected by husband without any social support. This will also cause hatred between wife and husband in the homes causing unnecessary tension and worse of all, separation where the husband chases away the woman from his home due to anger*…*”* (FGD _Female 20+_Adjumani_Nyumanzi_Settlement)
>
> *“…And the worse of all may lead to separation where the husband chases away the woman from his home…”* (FGD _Female 20+_Adjumani_Mirieyi_Settlement)

## Discussions

Gender dynamics plays a very important role in many patriarchal societies ranging from influence of health behavior to decision making on critical issues [22]. In this research gender dynamics was found to influence the decisions taken against contraceptive use by couples, low empowerment of women to use contraception and patriarchal dominance of men in making contraceptive choices[23]. Commercialization of the girl child through marriage and dowry payment is not new in many low- and middle-income countries [24]. This has had a push-on effect on the aspiration of couples to jointly agree to have as many girl children as possible in anticipation of monetary value through marriage and subsequent dowry. This research found that in the refugee settings participants viewed contraceptive use as a barrier to getting the many cows in form of bride price that comes with a girl child. Families that produced many girls were assured of big gains in form of dowry. Therefore, participants reaffirmed that couples do agree jointly [25] not to use contraception so as to have as many girls as possible to reap the benefits of dowry. Considering the above assertions there is need for the ministry of health in conjunction with non-governmental organizations to roll out health promotion awareness on the benefits of family planning and the need to respect the integrity of the girl child. Such a strategy may also be effective if men are also involved in critical issues regarding contraception.

Empowerment of women has always been a very important and critical issue to offer women a space to make decision on what impacts on their health [26]. Empowered women are likely to be independent and fearless to make a decision that addresses their needs which has been mistaken for disobedience in many patriarchal societies. This research found that women who are less empowered are unable to convince their husbands about the intention to use contraceptives. They reported that personal decision is only possible for women who are economically empowered and can sustain themselves and provide basic needs should the husband discover that they are using any contraceptives. Considering the above assertions, there is a need for government to roll out assertiveness training among women including running the training alongside economic empowerment projects for women [1]. Furthermore, it is also important that men who already view themselves as the custodians of the community and central to decision making are also included the women empowerment awareness programme. This will help them to appreciate the benefits of an empowered woman not only to the family but also to the community.

In most patriarchal societies, men have dominated all decisions that pertains to family matters [27]. This male dominance has further exacerbated the vulnerability of women and girls affecting their access to health services. In some circumstance for women to access certain health services they must first get approval from their partners [28]. Due to the subordinate situation of the refugee women, they continuously experience barriers to accessing contraceptives. This study revealed that women could not make their own decision to use contraceptive because of male influence against the idea. The community leaders who are predominantly men are fully aware of this and are also enjoying the monopoly of making decisions on what type of healthcare should be accessed by women. They acknowledged that men ‘are regarded as the custodians of the community and have a responsibility of taking important decisions for their communities [29], thus, any decision made by women is rejected. Drawing from the above findings, this study recommends a tailored community intervention to be designed together with community leaders, men and women to create awareness on the rights of every individual particularly right to health. This will help to address the subordinate positions of women that has affected their lives for a long period of time. In doing so, they may appreciate and steer the process of re considering the rules governing patriarchy while at the same time allow women to take part in decision making especially about their health needs.

Use of contraceptives is aimed at achieving pregnancy prevention. However, in some communities, it is perceived as a characteristic of promiscuity [30]; [31]; [32]. The different cultural norms that have along viewed contraceptive use as synonymous to promiscuity have led to many refugee women fearing to access the service [30]. This study revealed that many husbands and parents of young women believed that women who used contraception are promiscuous. They added that the community viewed contraceptive use as a cover up for women’s intentions to engage in sexual relationships with other men outside their marital life, [32]. Given these concerns, there is need to clearly draw a line between pregnancy prevention particularly for child spacing and the community views on risk sexual behaviour. Pre packed messages showing benefits of family planning to baby, mother, father, community and government as a whole should be considered. This should be related to the harnessing of the Demographic Dividend that shows how fertility reduction will enhance dependency ratio and lead to saving that will benefit the family to be able to invest.

Contraceptive use has for a long period been perceived differently by many communities across the world [33]. The various myths and misconceptions have impacted negatively on the uptake of family planning programs in many African countries [30]; [31]; [34]. Although many, scholars and practitioners have written and talked about these issues, some communities including the refugee populations still strongly believe in these myths related to ill health, poor pregnancy outcome and fertility challenges among others [35]. In this study, participants reported that contraceptive use led to children being born with deformities which has deterred some refugee women from making decision to use contraceptives. Owing to the above findings, this study suggests that these there is need for the central government through the Ministry of Health and key humanitarian stakeholders to raise awareness about family planning within communities to dispel these myths and misconceptions. The awareness can take the form of dialogue with the communities to allow for questions and answers. These dialogues will provide space for both women and men to build a more supportive environment to interact with providers in addressing these myths. This approach will help address the challenges, answer questions from the community.

Cultural norms and values have a direct impact on the uptake of health services thus the impact may be positive or negative [36]. In many communities across Africa women who use contraceptives are viewed as weak and fear to raise children which is against what a woman is expected in the community [29]. This research found that the research participants believed that women were married to have as many children as they could without any interference of contraceptives. Furthermore, women were expected to give birth until menopause and to have a big family. Due to these strong cultural norms, women who fail to have the desired number of children are subjected to disciplinary action like being chased out of the matrimonial home or beaten [29]. Considering the assertions above, this study therefore recommends that the Central Government through the Ministry of Health and humanitarian organization working in the refugee settlements roll out a health promotion campaign to enlighten community leaders and men in general on the cultural issues that impact on the uptake of women contraceptive use. Such an initiative can help in attitude change and increase the uptake of contraception among refugee communities.

Limited knowledge about contraceptive methods have been found as barrier for decision making in general [30]; [34];[37]. The population under study come from a community where sensitization on family planning was never a government priority. It is clear that when people do not know about a service or have limited knowledge about it, they would not seek to use the service, [25]. The participants in this study reported that contraceptive information is not widely known or disseminated in their settlements. Additionally, they acknowledged that there was lack of health promotion messages that promoted family planning back home in South Sudan. Therefore, it is important for humanitarian organisation working with refugees to equip them with information on contraception. This study further recommends that the central government through the Ministry of Health and Adjumani district should come up with appropriate ways of disseminating family planning messages in the refugee settlements by working with community leaders and community health workers in the community.

Reprisal following contraceptive use is one of the ways in which men can restrict the uptake of certain health care services by women for example sexual reproductive health desires [29]; [34]. Such reprisals may involve neglecting women by leaving them without any social support thereby making them vulnerable considering their socio-economic status in the community [37]. Most families have fallen apart due to disagreements on contraceptive use, impacting negatively on the lives of children and the women themselves due to intimate partner violence [38]. Persistent gender norms have affected uptake of health services amongst young women leading to fear [39]. This research found that women who went ahead and used contraceptives without approval of their husbands and/or family members received some reprisals in form beating or separation/divorce. The refugee women believed that use of contraceptives was safer when the husbands and family members were not aware. Considering the above fears, interpersonal communication should be designed for men, women, local leaders to uphold the dignity of women in relation seeking health services. The men should also be sensitized about the consequences of intimate partner violence if they were acting out of ignorance. This will help them to appreciate the law and its implication on intimate partner violence while accepting contraceptive use by their women.

## Implication for practice

There is need for a multidisciplinary approach to combatting barriers to contraception use among refugee communities. Professionals from different organizations and departments need to be educated and trained to understand the culture of refugee communities and effectively help in promoting access to contraception services. More importantly there is need for a robust health promotion programme targeting both men and women in the refugee communities. Such a health promotion initiative should also be supported and conducted by people within the refugee communities to enhance acceptability. More conversations are needed with refugee women and men to understand the best ways of caring for the women and tap into opportunities available to break the barriers to contraceptive use. Such engagement may be key to finding lasting solutions in ending opposition to contraceptive use among refugee communities. There is need to adopt locally appropriate approaches and interventions to deliver health promotion to contraceptive awareness and paying attention to concept of social justice and recognition of local knowledge and values.

### Limitation of the study

The research participants were drawn from one district in Uganda in this case Adjumani. Future research encompassing several districts hosting refugees in Uganda will be needed to enhance comparisons. Furthermore, the research study was limited to only a qualitative research paradigm. However, future studies using both qualitative and quantitative research paradigms will be needed to explore the research issues from different epistemological and ontological positions.

## Conclusion and recommendation

Several constrains negatively affected the use of contraception among South Sudanese refugee women living in Uganda. This calls for community friendly strategies to break down the barriers to contraception utilization among refugee women. Such strategies should include both men and women alongside the community leaders to enhance sustainability. Acknowledgement of the values, knowledge and ethos of the refugee communities should be recognized to build a formidable relationship with the community leaders while selling the benefits of family planning at community level. More importantly, robust policies supporting family planning among refugee communities are needed to ensure that the public is aware of the need to engage with the family planning services and improve the women s’ health.

## Data Availability

Data is available and will be available on request.

## Funding

### Grant ID

NICHE -UGA-288. This research was conducted with financial support from Nuffic grant through The International Institute of Social Studies Erasmus University, Rotterdam for Strengthening Education and Training Capacity in Sexual Reproductive Health & Rights in Uganda (SET-SRHR).

### Funding to authors

Funding was only availed to RA, the Principal Investigator through Nuffic grant through The International Institute of Social Studies Erasmus University, Rotterdam for Strengthening Education and Training Capacity in Sexual Reproductive Health & Rights in Uganda (SET-SRHR). The funders had to role in the study design, data collection and analysis, decision to publish or preparation of the manuscript.

## Acknowledgements

We are grateful to all refugee women who took part in this research study and the research assistants for data collection. Special thanks go to my supervisors for the continued support.

## Author Contributions

**Conceptualization:** Roselline Achola (RA), Christopher Garimoi Orach (CGO), Lynn Atuyambe (LA) and Elizabeth Nabiwemba (EN)

**Formal analysis**: RA, CGO and Mathew Nyashanu (MN)

**Funding acquisition**: Ra, CGO, LA and EN

**Investigation**: RA, CGO, LA, EN and MN

Methodology: RA, CGO, LA, EN and MN

Project administration: Olivia Nakisita; Software: RA and Tenywa Ronald

Supervision: RA, CGO, LA and EN

Validation: RA, CGO, LA, EN and MN

Visualization: RA

Writing – original draft: RA, CGO, MN: Writing – review & editing: RA, CGO, LA, EN and MN

